# Diazepam modulates hippocampal CA1 functional connectivity in people at clinical high-risk for psychosis

**DOI:** 10.1101/2024.12.20.24319330

**Authors:** Nicholas R. Livingston, Amanda Kiemes, Owen O’Daly, Samuel R. Knight, Paulina B. Lukow, Luke A. Jelen, Thomas J. Reilly, Aikaterini Dima, Maria Antonietta Nettis, Cecilia Casetta, Gabriel A. Devenyi, Thomas Spencer, Andrea De Micheli, Paolo Fusar-Poli, Anthony A. Grace, Steve C.R. Williams, Philip McGuire, M. Mallar Chakravarty, Alice Egerton, Gemma Modinos

## Abstract

**Background:** Preclinical evidence suggests that diazepam enhances hippocampal γ-aminobutyric acid (GABA) signalling and normalises a psychosis-relevant cortico-limbic-striatal circuit. Hippocampal network dysconnectivity, particularly from the CA1 subfield, is evident in people at clinical high-risk for psychosis (CHR-P), representing a potential treatment target. This study aimed to forward-translate this preclinical evidence.

**Methods:** In this randomised, double-blind, placebo-controlled study, 18 CHR-P individuals underwent resting-state functional magnetic resonance imaging twice, once following a 5mg dose of diazepam and once following a placebo. They were compared to 20 healthy controls (HC) who did not receive diazepam/placebo. Functional connectivity (FC) between the hippocampal CA1 subfield and the nucleus accumbens (NAc), amygdala, and ventromedial prefrontal cortex (vmPFC) was calculated. Mixed-effects models investigated the effect of group (CHR-P placebo/diazepam vs. HC) and condition (CHR-P diazepam vs. placebo) on CA1-to-region FC.

**Results:** In the placebo condition, CHR-P individuals showed significantly lower CA1-vmPFC (*Z*=3.17, *P*_FWE_=0.002) and CA1-NAc (*Z*=2.94, *P*_FWE_=0.005) FC compared to HC. In the diazepam compared to placebo condition, CA1-vmPFC FC was significantly increased (*Z*=4.13, *P*_FWE_=0.008) in CHR-P individuals, and both CA1-vmPFC and CA1-NAc FC were normalised to HC levels. In contrast, compared to HC, CA1-amygdala FC was significantly lower contralaterally and higher ipsilaterally in CHR-P individuals in both the placebo and diazepam conditions (lower: placebo *Z*=3.46, *P*_FWE_=0.002, diazepam *Z*=3.33, *P*_FWE_=0.003; higher: placebo *Z*=4.48, *P*_FWE_<0.001, diazepam *Z*=4.22, *P*_FWE_<0.001).

**Conclusions:** This study demonstrates that diazepam can partially restore hippocampal CA1 dysconnectivity in CHR-P individuals, suggesting that modulation of GABAergic function might be useful in the treatment of this clinical group.

## Introduction

Identifying novel pharmacological interventions to reduce symptom severity and prevent transition to psychosis in individuals at clinical high-risk for psychosis (CHR-P) is a significant unmet clinical need^1,2^. Current neurobiological theories of psychosis development identify the hippocampus as a central hub of pathophysiology^47,59,107^ and a promising pharmacological target^6^. Several neuroimaging studies in individuals at CHR-P have identified increased hippocampal cerebral blood flow/volume compared to healthy controls (HC)^3–5^. The cornu ammonis 1 (CA1) subfield is proposed to be the origin of hippocampal dysfunction in the CHR-P state, in terms of volume loss^7^ and hyperactivity^8,9^, which then spreads to the subiculum following psychosis onset^9^. The CA1 and subiculum have a high number of glutamatergic efferent projections^10^, and anterior projections innervate a cortico-limbic-striatal circuit encompassing the nucleus accumbens (NAc) of the striatum, amygdala, and the ventromedial prefrontal cortex (vmPFC)^11^. These regions are highly interconnected^11–19^ and are associated with positive, negative, and cognitive symptoms of schizophrenia, respectively^20–22^. Therefore, hippocampal dysfunction preceding the onset of psychosis may disrupt downstream cortico-limbic-striatal regions, contributing to circuit dysfunction and the emergence of psychosis^11^.

Circuit dysfunction can be investigated in terms of the functional connectivity (FC) between brain regions measured using resting-state functional magnetic resonance imaging (rs-fMRI)^23^. rs-fMRI studies have identified altered hippocampal FC with the cortico-limbic-striatal circuit in individuals with a first episode of psychosis or chronic schizophrenia compared to HC. More specifically, these studies reported lower hippocampal FC with the striatum^24–30^ and vmPFC^24,26,28,29,29,31–40^, and either lower^38,41^, higher^42^, or unaltered^42^ hippocampal FC to the amygdala. The pattern is less clear in subclinical psychosis spectrum individuals (although there are far fewer studies): lower hippocampal-striatal FC has been shown in healthy individuals with high schizotypy traits^43,44^, while both lower^25,34^ and normal^33,45,46^ hippocampal-striatal and hippocampal-PFC FC have been observed in individuals at CHR-P compared to HC. To our knowledge, no studies in CHR-P individuals have investigated hippocampal-amygdala FC, or FC alterations from specific hippocampal subfields to the cortico-limbic-striatal circuit. Given that hippocampal dysfunction may be localised to the CA1 subfield in the CHR-P stage^8^, alterations in FC may may not be present across the whole hippocampus.

GABAergic dysfunction has been proposed as a key mechanism underlying hippocampal hyperactivity in psychosis^47^. Studies in rats exposed to the mitotoxin methylazoxymethanol acetate (MAM) during neurodevelopment showed that reduced PV+ interneuron number in the hippocampus was associated with an increased firing rate of local excitatory neurons and excitatory/inhibitory imbalance^48^. This hyperactivity is found to drive functional alterations of downstream regions in MAM-treated rats, evidenced by experiments where chemical^48^ or pharmacological inactivation of the hippocampus (with a nonspecific GABA_A_-enhancing benzodiazepine^49^ or an α5-GABA_A_ specific compound^49,50^) normalised midbrain dopaminergic neuron firing. Furthermore, this mechanism is proposed to underlie the findings that repeated peripubertal diazepam administration in MAM-treated rats prevented the emergence of schizophrenia-related neurophysiological and behavioural phenotypes in adulthood. Such phenotypes included prevention of midbrain dopamine hyperactivity and hyperlocomotion response to amphetamine (positive symptoms), amygdala hyperactivity (negative symptoms), and PFC dysfunction (cognitive symptoms)^51–53^.

This preclinical evidence suggests that GABA-enhancing compounds may be an effective strategy for psychosis prevention by downregulating hippocampal hyperactivity and normalising downstream circuit dysfunction. In healthy individuals, prior rs-fMRI studies using an acute, non-sedating dose of a GABA-enhancing compound report increases in FC under benzodiazepine (or other GABA-enhancing drugs e.g., Z-drugs such as zopiclone/zolpidem) compared to placebo across the hippocampal-amygdala-PFC circuit^54^, the default mode network^55,56^, and a wider brain network including visual, auditory, sensorimotor, and prefrontal regions^57^. In CHR-P individuals, we recently demonstrated that an acute, non-sedating dose of diazepam normalised elevated hippocampal and subfield cerebral blood flow towards levels seen in healthy controls^58^. However, whether this is accompanied by a normalisation of the FC between the hippocampus and downstream cortico-limbic-striatal regions was not known.

Therefore, the current study examined the effects of an acute dose of diazepam vs. placebo on FC between the hippocampus and this cortico-limbic-striatal circuit in the same cohort of CHR-P individuals^58^. Each condition was also compared to HC data collected on the same scanner. We focussed on the CA1 subfield as a seed, given its proposed role in psychosis development^59^ and the substantial number of anatomical connections to output regions of interest (NAc, amygdala, and vmPFC^60,61^). On the basis of previous findings in hippocampal FC across the psychosis spectrum^16–26,29–32,34–36,41,43^, we hypothesised that individuals at CHR-P (in the placebo condition) would display lower CA1-NAc and CA1-vmPFC FC and altered CA1-amygdala FC compared to HC. Based on prior benzodiazepine challenge rs-fMRI studies in healthy individuals^50–53^, we hypothesised that a single dose of diazepam would increase CA1 FC within this circuit, to the extent that it would no longer differ from HC. For completeness, the following supplementary analyses were included: i) using the anterior hippocampus as a seed (given it is specifically the anterior portion of the CA1 implicated in psychosis development^8,9^) and ii) exploring broader effects of diazepam on CA1/anterior hippocampus FC with the rest of the brain.

## Methods and Materials

### Study design, participants, and procedure

This experimental medicine study was conducted at King’s College London. The study received ethical approval from the National Health Service UK Research Ethics Committee (18/LO/0618), and each participant gave written informed consent. While the study received ethical clearance as ‘not a Clinical Trial of an Investigational Medicinal Product’ by the EU directive 2001/20/EC, it was registered on clinicaltrials.gov (NCT06190483). Full study details, including inclusion/exclusion criteria, can be found in our recent publication describing the hippocampal cerebral blood flow findings in the same participants^58^. Briefly, this study used a randomised, double-blind, placebo-controlled, crossover design, whereby 24 antipsychotic-naïve individuals at CHR-P underwent MRI scanning on two occasions, once following a single oral dose of diazepam (5mg) and once following an oral placebo (50mg ascorbic acid). The diazepam/placebo capsule was administered 60 minutes before MRI scanning, and there was a minimum 3-week washout period between scans. Data from a group of 22 HC from a prior study (PSYAUD17/25) acquired with the same MRI scanner, scanning sequences, and acquisition parameters were used as a comparison group^65^.

### MRI acquisition

MRI data were acquired on a General Electric MR750 3.0T MR scanner with an 8-channel head coil at the Centre for Neuroimaging Sciences, KCL. A 3D T1-weighted scan was acquired using a SPGR sequence and rs-fMRI data was acquired using a multi-echo echo planar imaging sequence (full acquisition details in Supplementary Methods). During the rs-fMRI scan, participants were instructed to remain awake with their eyes open, while a fixation cross was displayed in the centre of the screen.

### Neuroimaging data processing

#### Preprocessing

The structural and rs-fMRI data were preprocessed using fMRIPrep (version 23.1.3)^66^, SPM12^67^, CONN^68^, and FSL^69^. Structural images from both sessions were corrected for intensity non-uniformity using N4, skull-stripped, segmented, and averaged across sessions to generate a singular participant structural image which was then normalised to MNI space (1mm^3^ resolution)^66^. For the rs-fMRI data, volume re-alignment and slice-timing correction parameters were calculated using the first echo and applied to all echoes^66^. Participants were excluded if they moved >3mm on any translation/rotation parameter or had a mean framewise displacement of >0.5mm, as advised by prior methodological investigations^70^. The three echoes in native space underwent TE-dependent ICA-based denoising and were optimally-combined using T2* weighted averaging via TEDANA^71^, before being normalised to MNI space (2mm^3^ resolution) with transformations generated during fMRIPrep (see supplementary materials for full boiler plate)^66^. The denoised, optimally combined, normalised functional data was then spatially smoothed in SPM12^67^ with a 6mm FWHM Gaussian kernel, and further denoised by removing white matter and CSF signal using the first 5 components of aCompCor, despiking, scrubbing, and band-pass filtering (0.008-0.09 Hz) in CONN^68^.

#### Generation of seed and region-of-interest masks

Hippocampal and subfield seed masks were generated for each participant from their preprocessed structural scan collected during their first scanning visit using the MAGeT Brain (multiple automatically generated templates of different brains) toolbox^72^ (see previous publication for further details^58^). Using all participants’ CA1 segmentations, study-specific left and right CA1 masks were generated by using majority vote (ANTs/2.5.0; Figure 1). ROI masks for the cortico-limbic-striatal circuit (NAc, amygdala, and vmPFC) were derived from Neurosynth (https://www.neurosynth.org/) using the search terms ‘nucleus accumbens’, ‘amygdala’, and ‘vmPFC’ (uniformity tests). The resulting images were thresholded, binarised, and dilated (MINC toolkit; https://bic-mni.github.io/).

**Figure 1.**
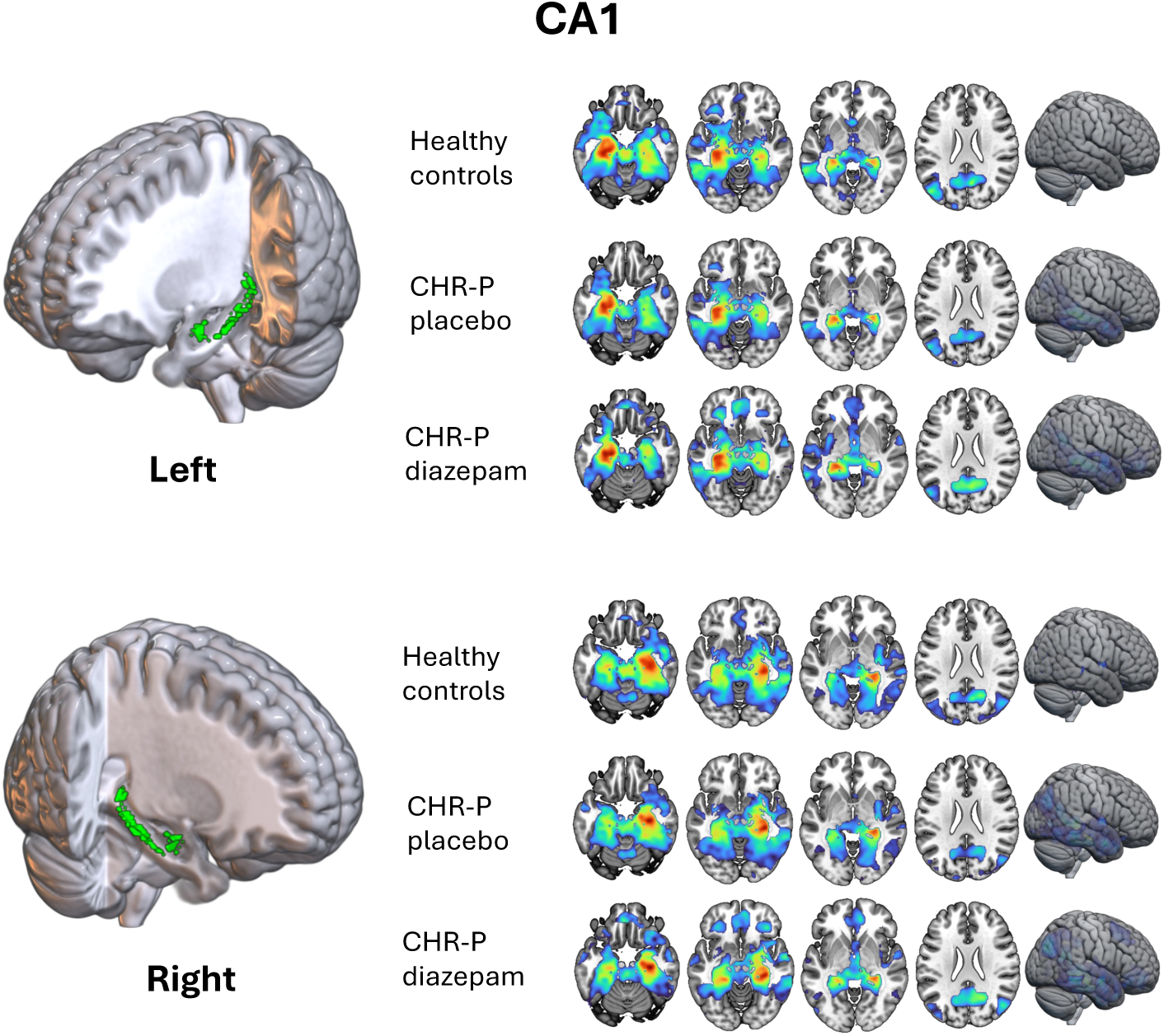
Within-group CA1-to-voxel functional connectivity. CA1-to-voxel functional connectivity networks averaged across each group independently (healthy controls, CHR-P placebo, and CHR-P diazepam) for the left and right CA1 subfield using study-specific mask (*Z* > 2.3, *P*_FWE_ < 0.05). *CHR-P: clinical high-risk for psychosis*

### Neuroimaging data analysis

To control for the number of models, FDR correction was performed on all FWE-corrected second-level analyses described below.

#### First- and second-level analysis

To generate participant-level seed-to-voxel *Z*-maps, the mean functional time series was extracted from the left and right CA1 and used in first-level analysis models as regressors of interest in FSL. These first-level seed-to-voxel *Z*-maps were then entered into second-level analysis models using FLAME-1 (FMRIB’s Local Analysis of Mixed Effects)^69^, which employs Bayesian modelling and a weighted least-squares approach to perform a mixed-effects analysis. FLAME-1 was chosen as mixed-effects modelling is optimal for within-subject designs (i.e., CHR-P diazepam vs. placebo) to account for within-subject correlations, and FLAME-1 is able to estimate different variances for different groups of subjects within a model, which is advantageous for unpaired two-sample comparison (i.e., CHR-P vs. HC)^73^. All models below use an FWE-corrected (*P*<0.05) threshold of *Z*>2.3. This threshold with FLAME-1 models has been shown to produce FWE rates lower than 5%, and is therefore similar to traditional FSL ordinary least square analyses using a threshold of *Z*>3.1^74^.

#### Within-group CA1 resting-state FC analyses

Before comparing differences between groups/conditions, we first validated within-group resting-state FC networks for the CA1 to the whole brain to ensure they matched expected networks (one-sample contrast for each group independently)^75^.

#### Group and condition seed-to-ROI analyses

To investigate the effect of group (CHR-P placebo/diazepam vs. HC) and condition (CHR-P diazepam vs. placebo) on FC differences between CA1 and cortico-limbic-striatal circuit regions, we conducted seed-to-ROI analysis. Second-level models were run per seed-to-ROI per hemisphere for each group/condition comparison using a small volume adjustment approach by applying a pre-threshold ROI mask generated from Neurosynth as described above. Models were run both contralaterally (e.g., left CA1 to the right amygdala) and ipsilaterally (e.g., left CA1 to the left amygdala), as disruptions to both have been found across the psychosis spectrum within this circuit^76^. Voxel-level thresholding was used (*Z>*2.3) for inference, which was FWE-corrected (*P*<0.05) for multiple comparisons. Again, this threshold has been demonstrated to be quite conservative when using voxel-level inference in FLAME-1 models^74^. For CHR-P placebo/diazepam vs. HC models, age (mean centred) and sex were added in as covariates of no interest. For CHR-P diazepam vs. placebo, change in pre-post scan fatigue score from the Bodily Symptoms Scale^77^ for each condition was included as a covariate of no interest to control for drug effects of sedation/fatigue.

#### Supplementary / exploratory analyses

For completeness, supplementary analyses explored the effect of group/condition on FC between 1) anterior hippocampus and the cortico-limbic-striatal circuit (seed-to-ROI), and 2) hippocampal seeds (CA1 and anterior hippocampus) and the rest of the brain on a voxel-wise basis. Anterior hippocampal masks were generated by masking the study-specific averaged whole hippocampus segmentation with a hippocampus head mask derived from the Allen human reference atlas^78^, then thresholded, binarized, and dilated. Identical second-level models were run as described above, and for the seed-to-voxel analyses an inclusive grey matter mask was used during the pre-threshold masking.

## Results

### Demographics and Clinical Assessments

Following data quality checks, 6 CHR-P participants were excluded (n=2 missing rs-fMRI data, n=1 poor data quality, n=3 excessive motion), along with 2 HC participants (n=1 missing rs-fMRI data, n=1 poor data quality). This resulted in a final sample of 18 CHR-P and 20 HC for analyses. Participant details can be found in Table 1.

**Table1.** Participant demographic information, clinical characteristics, head motion parameters and fatigue scores.

| | CHR-P<br>(n = 18) | HC<br>(n = 20) | Comparison | *t*/$\chi^2$ | *p* |
| --- | --- | --- | --- | --- | --- |
| **Demographic** |  |  |  |  |  |
| Age (years; mean ±SD) | 24.1 ±4.6 | 26.5 ±5.1 | CHR-P vs. HC | 1.5 | 0.136 |
| Sex (male/female; n) | 5/13 | 9/11 | CHR-P vs. HC | 1.2 | 0.272 |
| Ethnicity (n) |  |  | CHR-P vs. HC | 11.2 | **0.024** |
| Asian | 1 | 6 | - | - | - |
| Black | 4 | 0 | - | - | - |
| Mixed or multiple | 2 | 0 | - | - | - |
| Other | 1 | 0 | - | - | - |
| White | 10 | 14 | - | - | - |
| IQ (WAIS-III short version<sup>81</sup>; mean ±SD) | 96.0 ±22.1 | 122.9 ±13.9 | CHR-P vs. HC | 4.4 | **<0.001** |
| Current daily cigarette use, n (%) | 4 (22) | 2 (10) | CHR-P vs. HC | 1.1 | 0.302 |
| Current alcohol use, n (%) | 14 (77) | 18 (90) | CHR-P vs. HC | 1.1 | 0.302 |
| Current cannabis use, n (%) | 5 (28) | 3 (15) | CHR-P vs. HC | 1.8 | 0.181 |
| **Clinical characteristics** |  |  |  |  |  |
| CAARMS<sup>83</sup> score (mean ±SD) | | | | | |
| Positive symptoms | 47.5 ±12.9 | NA | - | - | - |
| Negative symptoms (n=21) | 29.5 ±25.3 | NA | - | - | - |
| Total (n=21) | 77.9 ±28.2 | NA | - | - | - |
| Global functioning score<sup>85</sup> (mean ±SD) | | | | | |
| Social | 6.3 ±1.5 | NA | - | - | - |
| Role | 6.1 ±1.7 | NA | - | - | - |
| Hamilton scale score (mean ±SD) | | | | | |
| Anxiety<sup>87</sup> (n=22) | 17.6 ±9.3 | NA | - | - | - |
| Depression<sup>89</sup> (n=21) | 13.5 ±6.9 | NA | - | - | - |
| Current antidepressant medication, n (%) | 7 (38) | NA | - | - | - |
| Current or prior antipsychotic medication, n (%) | 0 (0) | NA | - | - | - |
| Current benzodiazepine/hypnotic medication, n (%) | 0 (0) | NA | - | - | - |
| **Head motion** |  |  |  |  |  |
| Fractional displacement in mm (mean ±SD) | | | | | |
| Total group | 0.148 ±0.08 | 0.146 ±0.07 | CHR-P vs. HC | 0.05 | 0.957 |
| Placebo condition | 0.156 ±0.10 | - | CHR-P placebo vs. HC | 0.36 | 0.724 |
| Diazepam condition | 0.139 ±0.07 | - | CHR-P diazepam vs. HC | -0.35 | 0.722 |
|  | - | - | CHR-P diazepam vs. placebo | 0.62 | 0.544 |
| **Bodily Symptoms Scale** |  |  |  |  |  |
| Fatigue scores post-scan (mean ±SD) | | | | | |
| Placebo condition | 0.944 ±0.93 | - | CHR-P diazepam vs. placebo | 1.51 | 0.148 |
| Diazepam condition | 1.38 ±1.58 | - | - | - | - |

CAARMS: comprehensive assessment of at-risk mental states; CHR-P: clinical high-risk for psychosis; HC: healthy control; IQ: intelligent quotient; WAIS: Weschler adult intelligence scale

CHR-P individuals had a significantly lower IQ (as is often observed in this clinical population^79^) and differed in terms of ethnicity compared to the HC group (which was driven by a high proportion of white ethnicity in the HC group). There were no significant differences in head motion parameters or change between pre- and post-scan Bodily Symptom Scale^77^ scores between the placebo and diazepam conditions.

### Resting-State Functional Connectivity

#### Within-group CA1 resting-state FC

Within each group/condition, as expected^75^, the CA1 showed significant FC with the rest of the hippocampus, extending to the temporal lobe, amygdala, precuneus, posterior cingulate cortex, mPFC, and parieto-occipital regions (*Z*>2.3, *P*_FWE_<0.05; Figure 1).

#### CA1-to-ROI

Compared to HC, individuals at CHR-P in the placebo condition showed significantly lower FC between the left CA1 and the right NAc (Figure 2A, Table 2), and between the right CA1 and the left NAc, left amygdala, and left vmPFC (Figure 2B, Table 2).

**Figure 2.**
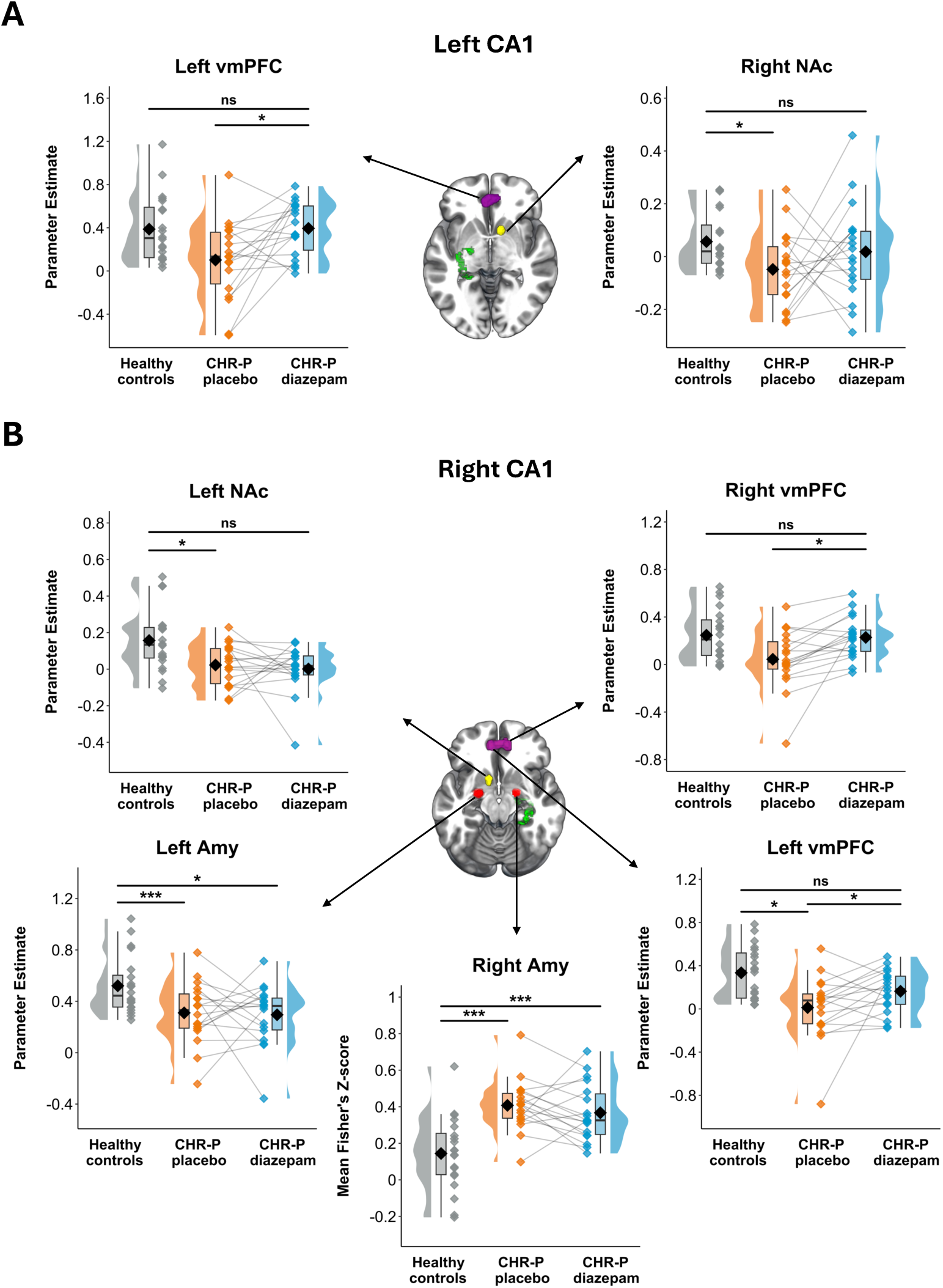
Region-of-interest functional connectivity results for the CA1. Parameter estimates of functional connectivity strength between left **(A)** and right **(B)** CA1 and output regions (nucleus accumbens, amygdala, and ventromedial prefrontal cortex) displayed for healthy controls and individuals at clinical high-risk for psychosis (in the placebo and diazepam conditions) at peak coordinate of significant effect of group/condition (*Z* > 2.3, *P*_FWE_ < 0.05). CA1 (green), amygdala (red), nucleus accumbens (yellow), and vmPFC (purple) are visualised on the brain using masks. *CHR-P clinical high-risk for psychosis; Amy: amygdala; NAc: nucleus accumbens; vmPFC: ventromedial prefrontal cortex; *** < 0.001; * < 0.05, ns not significant*

**Table 2.**
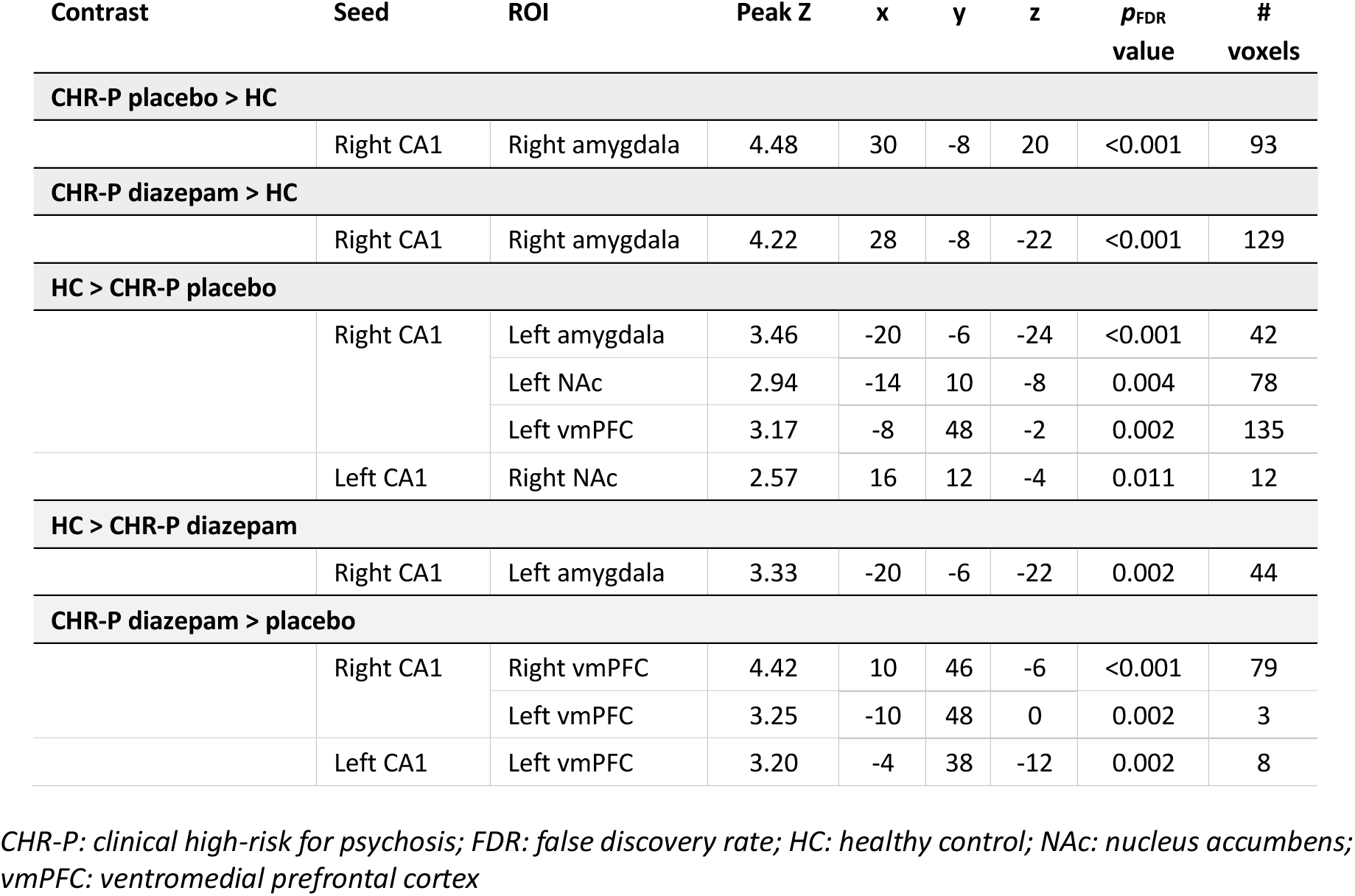
Summary sta;s;cs of region-of-interest functional connectivity results for the CA1.

| Contrast | Seed | ROI | Peak Z | x | y | z | $p_{FDR}$<br>value | #<br>voxels |
| --- | --- | --- | --- | --- | --- | --- | --- | --- |
| **CHR-P placebo > HC** |  |  |  |  |  |  |  |  |
|  | Right CA1 | Right amygdala | 4.48 | 30 | -8 | 20 | <0.001 | 93 |
| **CHR-P diazepam > HC** |  |  |  |  |  |  |  |  |
|  | Right CA1 | Right amygdala | 4.22 | 28 | -8 | -22 | <0.001 | 129 |
| **HC > CHR-P placebo** |  |  |  |  |  |  |  |  |
|  | Right CA1 | Left amygdala | 3.46 | -20 | -6 | -24 | <0.001 | 42 |
|  |  | Left NAc | 2.94 | -14 | 10 | -8 | 0.004 | 78 |
|  |  | Left vmPFC | 3.17 | -8 | 48 | -2 | 0.002 | 135 |
|  | Left CA1 | Right NAc | 2.57 | 16 | 12 | -4 | 0.011 | 12 |
| **HC > CHR-P diazepam** |  |  |  |  |  |  |  |  |
|  | Right CA1 | Left amygdala | 3.33 | -20 | -6 | -22 | 0.002 | 44 |
| **CHR-P diazepam > placebo** |  |  |  |  |  |  |  |  |
|  | Right CA1 | Right vmPFC | 4.42 | 10 | 46 | -6 | <0.001 | 79 |
|  |  | Left vmPFC | 3.25 | -10 | 48 | 0 | 0.002 | 3 |
|  | Left CA1 | Left vmPFC | 3.20 | -4 | 38 | -12 | 0.002 | 8 |

*CHR-P: clinical high-risk for psychosis; FDR: false discovery rate; HC: healthy control; NAc: nucleus accumbens; vmPFC: ventromedial prefrontal cortex*

Additionally, the right CA1 showed higher FC to the right amygdala (Figure 2B, Table 2). In the diazepam condition, these differences observed in the placebo condition compared to HC were ameliorated (no significant difference), apart from the right CA1 to left and right amygdala, which still showed significantly lower and higher FC compared to HC, respectively (Figure 2B, Table 2). We observed a significant drug effect on CA1-vmPFC FC, where diazepam (compared to placebo) significantly increased the FC strength from the left CA1 to left vmPFC and right CA1 to bilateral vmPFC (Figure 2, Table 2).

#### Supplementary / exploratory analyses

At the whole-brain level, compared to HC, individuals at CHR-P in the placebo condition showed significantly higher FC between the right CA1 and a right medial temporal network including the hippocampus, insula, and inferior/medial temporal gyri (Figure 3A, Supplementary Table 1). Conversely, lower FC was observed between the right CA1 and a left medial temporal network that extended to include key regions of the default mode network (bilateral mPFC, anterior cingulate cortex, and posterior cingulate cortex). In the diazepam condition, higher FC between right CA1 and a right medial temporal lobe network was also observed compared to HC, and additionally extended to parieto-occipital regions such as the angular gyrus (Figure 3B, Supplementary Table 1). When comparing CHR-P diazepam vs. placebo conditions directly, no significant differences in whole-brain FC were observed for either the right or left CA1. Finally, there were no significant differences between groups (CHR-P placebo/diazepam vs. HC) or conditions (CHR-P diazepam vs. placebo) in FC strength using the anterior hippocampus as a seed on a ROI or whole-brain level.

**Figure 3.**
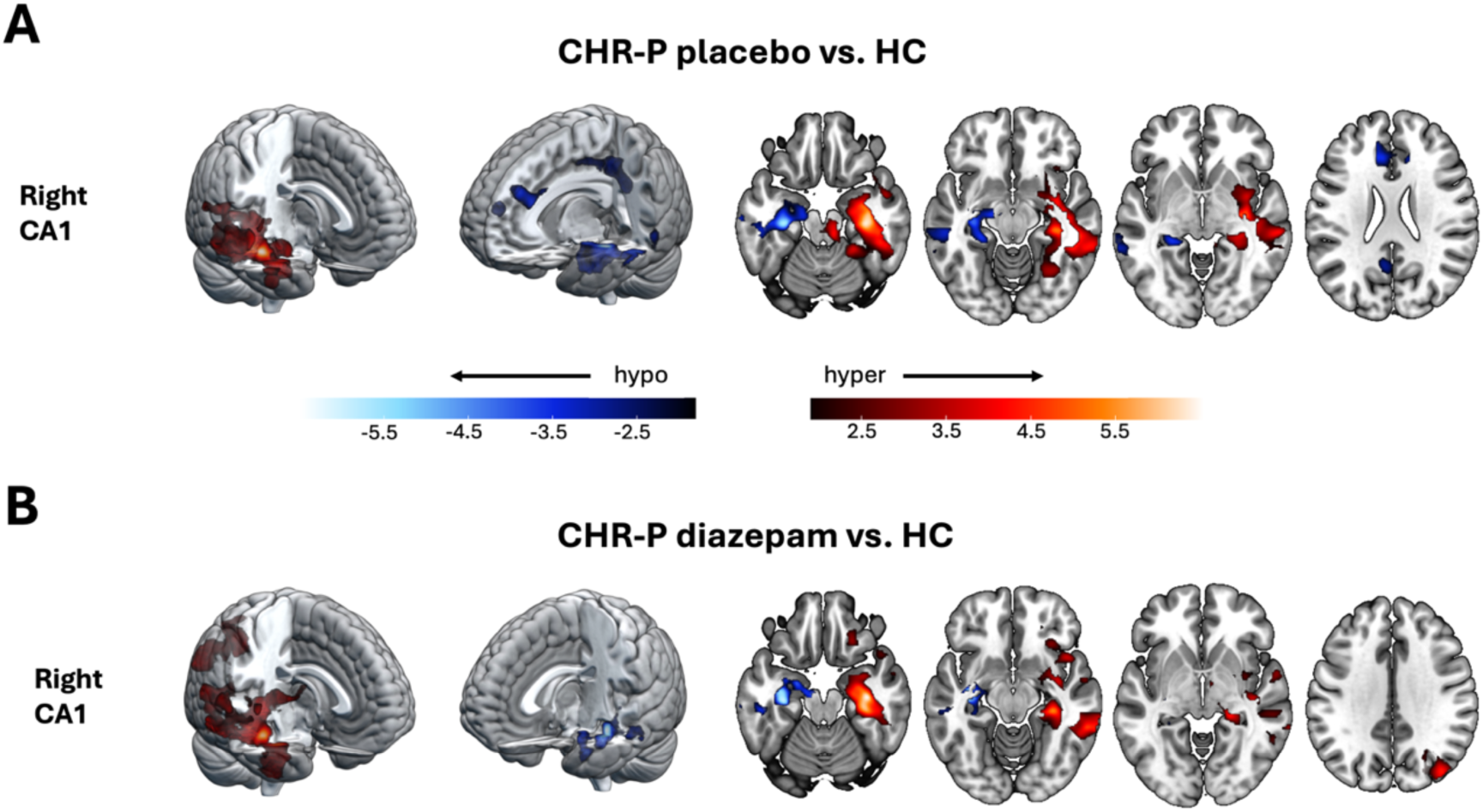
Voxel-wise whole-brain functional connectivity results for the CA1. Significant clusters showing differences (Z > 2.3, *P*_FWE_ < 0.05) in functional connectivity between healthy controls and CHR-P placebo **(A)** and CHR-P diazepam **(B)** for the CA1. Areas showing functional hyperconnectivity (CHR-P placebo/diazepam > HC) are displayed in red colourbar, whilst areas displaying functional hypoconnectivity are displayed in blue. N.B., no significant differences were found for the anterior hippocampus, nor for any of the regions (CA1 or anterior hippocampus) when contrasting CHR-P diazepam vs. CHR-P placebo. *CHR-P clinical high-risk for psychosis; HC healthy controls*

## Discussion

The main finding of the current study was that a single, non-sedating dose of the GABA-enhancing drug diazepam partially normalised CA1 dysconnectivity to a cortico-limbic-striatal circuit in individuals at CHR-P. More specifically, CHR-P individuals in the placebo condition (compared to HC) showed lower CA1-vmPFC and CA1-NAc FC. Diazepam significantly increased CA1-vmPFC FC compared to placebo, and the lower CA1-vmPFC and CA1-NAc FC observed in the placebo condition was normalised to HC levels. We observed more complex results for CA1-amygdala FC, as CHR-P individuals in the placebo condition showed lower and higher FC compared to HC, which were still present in the diazepam condition. Previously, we demonstrated that diazepam normalised increased hippocampal and subfield regional cerebral blood flow in the same CHR-P individuals, and here we extend this work by showing that diazepam can also partially normalise CA1 dysconnectivity to a downstream circuit. Taken together, these results indicate that GABA-enhancing compounds can rescue brain function in a psychosis-relevant circuit in CHR-P individuals, and therefore show promise as a novel treatment strategy for clinical intervention in this group.

Our finding of lower CA1-vmPFC and CA1-NAc FC contralaterally (but normal FC ipsilaterally) in CHR-P individuals in the placebo condition (vs. HC) is consistent with prior rs-fMRI reports of *subtle* dysconnectivity in sub-clinical psychosis populations^25,33,34,41,43–46^. In contrast, studies in first-episode and chronic schizophrenia samples *consistently* report lower FC between these regions^24–28,28–40^. This may suggest that in psychosis vulnerability stages, as hippocampal hyperactivity begins to drive glutamatergic input to the cortico-limbic-striatal circuit^11,20,21^, there is preserved temporal coherence (i.e., FC) between the hippocampus and these regions. As CHR-P symptoms persist, hippocampal hyperactivity and dysrhythmia may lead to uncoupling between the hippocampus and downstream circuitry, which may further deteriorate following the onset of psychosis. For example, experiments in MAM-treated rats demonstrated that NAc hyperactivity due to hippocampal dysfunction, drives a striatal-midbrain circuit loop^48^ which increases phasic dopamine efflux in the NAc itself^80^. Importantly, this increase in phasic dopamine can potentiate the hippocampal drive on the NAc^82^, which may result in reduced hippocampal-NAc FC. This inverse relationship of hippocampal hyperactivity and reduced hippocampal-striatal FC has been observed previously in individuals at CHR-P, as higher hippocampal glutamate levels (indicative of hyperactivity) was associated with reduced hippocampal-striatal FC^45^. In accordance with this, reduced CA1-NAc FC was the most robust finding in our sample of individuals at CHR-P (i.e., it was observed bilaterally in the CA1), in whom we have previously demonstrated hippocampal hyperactivity^58^. Beyond illness chronicity, the more pronounced reductions observed in hippocampal FC in individuals with psychotic disorders compared to those at CHR-P might be related to antipsychotic treatment. For instance, we observed reductions in right, but not left, CA1-vmPFC FC in our sample of antipsychotic-naïve individuals at CHR-P compared to the more robust observations in antipsychotic-treated individuals with schizophrenia^26,28,31–35^. Whilst cognitive symptoms which are present in the prodrome may worsen following the onset of psychosis^84,86,88^, chronic antipsychotic treatment may also play a role in further cognitive impairment related to hippocampal-PFC FC uncoupling^90,91^.

We found both higher and lower CA1-amygdala FC in individuals at CHR-P in the placebo condition compared to HC. Prior rs-fMRI studies have found lower^38,41^ and normal^42^ hippocampal-amygdala FC in individuals with psychotic disorders. However, hippocampal-amygdala FC was increased in people with schizophrenia with paranoia vs. no paranoia^42^, and higher hippocampal-amygdala-PFC FC was associated with higher fear/anxiety in individuals with early psychosis^92^. Whilst amygdala dysfunction is associated with negative symptoms of schizophrenia^21^, it is also implicated in clinically distinct comorbid anxiety/mood disorders, which are more common in those at CHR-P^93,94^. This increased affective component might explain the higher hippocampal-amygdala FC observed in our sample of individuals at CHR-P compared to HC. Furthermore, the findings in our study appeared hemisphere dependent (i.e., the right CA1 showed increased FC to the right amygdala and decreased FC to the left amygdala). This was also observed at the whole-brain level, whereby the right CA1 showed hyperconnectivity with a right medial temporal network, including the amygdala, but hypoconnectivity with a left hippocampal network and frontal regions of the left default mode network. Increased hippocampal FC with the medial temporal lobe has been observed previously in the psychosis spectrum^26,95,96^, and could therefore be driving hyperconnectivity to the amygdala given the close proximity and number of bidirectional connections^97^. Furthermore, this pattern of intra-hemispheric hyperconnectivity and inter-hemispheric hypoconnectivity has been found previously in individuals with psychotic disorders, indicating increased local network segregation and decreased remote network integration^98^.

The main effect of diazepam vs. placebo in CHR-P individuals on CA1 FC to the cortico-limbic-striatal network was a bilateral increase in CA1-vmPFC FC. Furthermore, all decreases in CA1-vmPFC and CA1-NAc FC in CHR-P individuals in the placebo condition compared to HC were not present in the diazepam condition. The general direction of the drug effect (that is, increasing FC) is in line with our predictions and with prior pharmacological rs-fMRI studies using acute doses of GABA-enhancing drugs in healthy individuals^54–57,64^. GABA-enhancing drugs, such as diazepam, are positive allosteric modulators of the GABA_A_ receptors via the benzodiazepine site^99^. Most commonly, benzodiazepine binding leads to increased hyperpolarisation of post-synaptic glutamatergic pyramidal cells^99^, reducing their activity^100^. The mechanism by which inhibition of neural activity in one brain region can result in increased FC to another has been recently elucidated by chemogenetic fMRI study in mice. Rocchi and colleagues^101^ demonstrated that either acute or chronic inhibition of the PFC led to increases in FC with direct thalamo-cortical output regions. The spiking activity was reduced but became more rhythmic and phase-locked to low-frequency oscillatory rhythms, leading to an increase in FC with connecting regions. Therefore, through this mechanism, it is likely that downregulation of hippocampal hyperactivity under diazepam (which we have demonstrated previously in this sample) led to increases in FC with connecting output regions.

Interestingly, the effect of diazepam on CA1-vmPFC FC showed the least inter-individual differences between people at CHR-P, whilst the effects in the amygdala and NAc were more varied. This may be due to the fact that the vmPFC, similar to the hippocampus, contains a high number of benzodiazepine receptors^102^. Consequentially, similar local effects on neural activity in the hippocampus and vmPFC might have also contributed to a more robust increase in temporal coherence between them. Increases in hippocampal-PFC FC under benzodiazepine vs. placebo have previously been reported^54^, along with increases in FC to somatosensory and occipital regions^103,104^ which also have high number of benzodiazepine binding sites^102^. Furthermore, as noted earlier, the largest alterations in hippocampal FC observed in individuals at CHR-P in the placebo condition compared to HC were with the amygdala. This suggests that hippocampal-amygdala FC was the most perturbed out of the cortico-limbic-striatal regions. Given the proposed role of the amygdala in the initiation of hippocampal hyperactivity^105^ and PV+ interneuron loss^106^, and the high number of connections between these regions^97^, a single dose of diazepam may not have been sufficient to regulate altered hippocampal-amygdala FC in individuals at CHR-P. In support of this, benzodiazepines have been shown to either increase^54^ or decrease^55^ hippocampal-amygdala FC in healthy individuals. This suggests the pharmacological effects of GABA-enhancing compounds on this circuity are inherently complex, without the presence of potential alterations to the GABAergic system in individuals at CHR-P.

Finally, we found no differences in FC strength between groups or drug conditions for the anterior hippocampus to the cortico-limbic-striatal circuit. This was unexpected, based on preclinical evidence^22^ and current theories about the pathophysiology of psychosis^47,59,107^. However, the anterior hippocampus contains subfields beyond the CA1 and subiculum, such as the CA2/3, which largely only have intra-hippocampal projections^108^. Therefore, inclusion of this signal may increase noise, making it difficult to detect subtle FC alterations between the anterior hippocampus and the cortico-limbic-striatal circuit within individuals at CHR-P. In line with this, whilst preclinical evidence focuses on the anterior hippocampus, it specifically identifies the anterior CA1 as the site of dysfunction^109^.

This study had several strengths. We used a gold standard randomised, double-blind, placebo-controlled, crossover study design in a sample of antipsychotic naïve individuals at CHR-P. The hippocampus and CA1 subfield were segmented with a high degree of accuracy using novel computational methods^72^, allowing the generation of study-specific hippocampal and subfield masks. We acquired rs-fMRI data using an advanced multi-echo sequence, allowing robust data cleaning and removal of non-physiological noise with advanced methodological techniques such as TEDANA^71^. This led to high quality data as within-group/condition resting-state FC networks for the CA1 to the rest of the brain replicated those found previously^75^. We were able to contextualise baseline differences and direction of drug effects in the CHR-P group by comparing them with data from a HC group. Finally, we used advanced statistical mixed-effects modelling, which is optimal for examining both inter-group differences without assuming uniform variance and also for investigating within-subject effects^73^. This study also had some limitations. Our sample size of CHR-P individuals was reduced from 24 down to 18 after quality control, but retrospective power analysis demonstrated that the diazepam vs. placebo analyses (mean Cohen’s *d*=0.83) had an achieved power of 91%. Additionally, this study was not powered to investigate relationship between FC alterations and symptoms, which would require a much larger CHR-P sample. Due to limitations with the resolution of rs-fMRI, we were not able to investigate differences in FC from specifically the anterior CA1 which is of particular relevance for psychosis^22^.

In conclusion, this study provides evidence that a single dose of a non-specific GABA-enhancing drug, such as diazepam, can normalise CA1 FC alterations with the vmPFC and NAc in individuals at CHR-P. Conversely, CA1-amygdala FC was greatly perturbed in people at CHR-P under placebo compared to HC and was largely unaffected by diazepam challenge. Given this mechanistic evidence, future research is warranted with extended treatment durations to link these neurobiological changes to symptoms and clinical outcomes, including psychosis prevention.

## Data Availability

Data produced in the present study are available upon reasonable request to the authors

## Acknowledgments

This research was funded by the Wellcome Trust and The Royal Society (202397/Z/16/Z to GM) and the National Institute for Health and Care Research (NIHR) Maudsley Biomedical Research Centre (BRC). The views expressed are those of the authors and not necessarily those of the Welcome Trust, NIHR or the Department of Health and Social Care. For the purpose of open access, the author has applied a CC-BY public copyright licence to any Author Accepted Manuscript version arising from this submission. NRL was funded by an MRC DTP PhD studentship at the time of data collection and analysis. PBL was in receipt of a PhD studentship funded by the NIHR Maudsley BRC at the time of data collection. OO is funded by the Maudsley BRC. LAJ was supported by an MRC Clinical Research Training Fellowship (MR/T028084/1) at the time of data collection. TJR is supported by an MRC Clinical Research Training Fellowship (MR/W015943/1). PFP is supported by the European Union funding within the MUR PNRR Extended Partnership initiative on Neuroscience and Neuropharmacology (Project no. PE00000006 CUP H93C22000660006 “MNESYS, A multiscale integrated approach to the study of the nervous system in health and disease”). AAG received funding from USPHS NIMH MH57440. MMC receives salary support from the Fonds de Recherche Québec – Santé and from a James McGill Professorship. MMC also receives research support from Canadian Institutes of Health Research, Natural Sciences and Engineering Research Council – Canada, McGill University’s Health Brains for Health Lives (a Canada Research Excellence Fund Iniative), and TRIDENT (a New Frontiers in Research Fund program).

