## Supplementary Table 1 for "Diazepam modulates hippocampal CA1 functional connectivity in people at clinical high-risk for psychosis"

**Supplemental methods**

**MRI acquisition details**

T1-weighted scan was acquired using a SPGR sequence (TR/TE/TI=7.31ms/3.02ms/400ms, flip angle=11°, FoV=270, voxel-size=1.05x1.05x1.20mm^2^, slice thickness=1.2 mm, 196 slices) and rs-fMRI data was acquired using a multi-echo echo planar imaging sequence (TR/TE=2500ms/12, 28, 44ms; flip angle=80°; FoV=240mm; voxel-size=3.75x3.75x4.0mm^2^; slice thickness=3mm; 32 slices; 192 volumes).

**fMRIPrep boilerplate output**

Results included in this manuscript come from preprocessing

performed using *fMRIPrep* 23.1.3

(@fmriprep1; @fmriprep2; RRID:SCR_016216),

which is based on *Nipype* 1.8.6

(@nipype1; @nipype2; RRID:SCR_002502).

Anatomical data preprocessing

: A total of 2 T1-weighted (T1w) images were found within the input

BIDS dataset.

All of them were corrected for intensity non-uniformity (INU)

with `N4BiasFieldCorrection` [@n4], distributed with ANTs (version unknown) [@ants, RRID:SCR_004757].

The T1w-reference was then skull-stripped with a *Nipype* implementation of

the `antsBrainExtraction.sh` workflow (from ANTs), using OASIS30ANTs

as target template.

Brain tissue segmentation of cerebrospinal fluid (CSF),

white-matter (WM) and gray-matter (GM) was performed on

the brain-extracted T1w using `fast` [FSL (version unknown), RRID:SCR_002823,

@fsl_fast].

An anatomical T1w-reference map was computed after registration of

2 T1w images (after INU-correction) using

`mri_robust_template` [FreeSurfer 7.3.2, @fs_template].

Brain surfaces were reconstructed using `recon-all` [FreeSurfer 7.3.2,

RRID:SCR_001847, @fs_reconall], and the brain mask estimated

previously was refined with a custom variation of the method to reconcile

ANTs-derived and FreeSurfer-derived segmentations of the cortical

gray-matter of Mindboggle [RRID:SCR_002438, @mindboggle].

Volume-based spatial normalization to one standard space (MNI152NLin2009cAsym) was performed through

nonlinear registration with `antsRegistration` (ANTs (version unknown)),

using brain-extracted versions of both T1w reference and the T1w template.

The following template was were selected for spatial normalization

and accessed with *TemplateFlow* [23.0.0, @templateflow]:

*ICBM 152 Nonlinear Asymmetrical template version 2009c* [@mni152nlin2009casym, RRID:SCR_008796; TemplateFlow ID: MNI152NLin2009cAsym].

Functional data preprocessing

: For each of the 2 BOLD runs found per subject (across all

tasks and sessions), the following preprocessing was performed.

First, a reference volume and its skull-stripped version were generated

from the shortest echo of the BOLD run using a custom

methodology of *fMRIPrep*.

Head-motion parameters with respect to the BOLD reference

(transformation matrices, and six corresponding rotation and translation

parameters) are estimated before any spatiotemporal filtering using

`mcflirt` [FSL , @mcflirt].

BOLD runs were slice-time corrected to 1.21s (0.5 of slice acquisition range

0s-2.42s) using `3dTshift` from AFNI [@afni, RRID:SCR_005927].

The BOLD time-series (including slice-timing correction when applied)

were resampled onto their original, native space by applying

the transforms to correct for head-motion.

These resampled BOLD time-series will be referred to as *preprocessed

BOLD in original space*, or just *preprocessed BOLD*.

A T2★ map was estimated from the preprocessed EPI echoes, by voxel-wise fitting

the maximal number of echoes with reliable signal in that voxel to a monoexponential signal

decay model with nonlinear regression. The T2★/S0 estimates from a log-linear regression fit were used for initial values.

The calculated T2★ map was then used to optimally combine preprocessed BOLD across

echoes following the method described in [@posse_t2s].

The optimally combined time series was carried forward as the *preprocessed BOLD*.

The BOLD reference was then co-registered to the T1w reference using

`bbregister` (FreeSurfer) which implements boundary-based registration [@bbr].

Co-registration was configured with six degrees of freedom.

First, a reference volume and its skull-stripped version were generated

using a custom

methodology of *fMRIPrep*.

Several confounding time-series were calculated based on the

*preprocessed BOLD*: framewise displacement (FD), DVARS and

three region-wise global signals.

FD was computed using two formulations following Power (absolute sum of

relative motions, @power_fd_dvars) and Jenkinson (relative root mean square

displacement between affines, @mcflirt).

FD and DVARS are calculated for each functional run, both using their

implementations in *Nipype* [following the definitions by @power_fd_dvars].

The three global signals are extracted within the CSF, the WM, and

the whole-brain masks.

Additionally, a set of physiological regressors were extracted to

allow for component-based noise correction [*CompCor*, @compcor].

Principal components are estimated after high-pass filtering the

*preprocessed BOLD* time-series (using a discrete cosine filter with

128s cut-off) for the two *CompCor* variants: temporal (tCompCor)

and anatomical (aCompCor).

tCompCor components are then calculated from the top 2% variable

voxels within the brain mask.

For aCompCor, three probabilistic masks (CSF, WM and combined CSF+WM)

are generated in anatomical space.

The implementation differs from that of Behzadi et al. in that instead

of eroding the masks by 2 pixels on BOLD space, a mask of pixels that

likely contain a volume fraction of GM is subtracted from the aCompCor masks.

This mask is obtained by dilating a GM mask extracted from the FreeSurfer's *aseg* segmentation, and it ensures components are not extracted

from voxels containing a minimal fraction of GM.

Finally, these masks are resampled into BOLD space and binarized by

thresholding at 0.99 (as in the original implementation).

Components are also calculated separately within the WM and CSF masks.

For each CompCor decomposition, the *k* components with the largest singular

values are retained, such that the retained components' time series are

sufficient to explain 50 percent of variance across the nuisance mask (CSF,

WM, combined, or temporal). The remaining components are dropped from

consideration.

The head-motion estimates calculated in the correction step were also

placed within the corresponding confounds file.

The confound time series derived from head motion estimates and global

signals were expanded with the inclusion of temporal derivatives and

quadratic terms for each [@confounds_satterthwaite_2013].

Frames that exceeded a threshold of 0.5 mm FD or

1.5 standardized DVARS were annotated as motion outliers.

Additional nuisance timeseries are calculated by means of principal components

analysis of the signal found within a thin band (*crown*) of voxels around

the edge of the brain, as proposed by [@patriat_improved_2017].

The BOLD time-series were resampled into standard space,

generating a *preprocessed BOLD run in MNI152NLin2009cAsym space*.

First, a reference volume and its skull-stripped version were generated

using a custom

methodology of *fMRIPrep*.

All resamplings can be performed with *a single interpolation

step* by composing all the pertinent transformations (i.e. head-motion

transform matrices, susceptibility distortion correction when available,

and co-registrations to anatomical and output spaces).

Gridded (volumetric) resamplings were performed using `antsApplyTransforms` (ANTs),

configured with Lanczos interpolation to minimize the smoothing

effects of other kernels [@lanczos].

Non-gridded (surface) resamplings were performed using `mri_vol2surf`

(FreeSurfer).

Many internal operations of *fMRIPrep* use

*Nilearn* 0.10.1 [@nilearn, RRID:SCR_001362],

mostly within the functional processing workflow.

For more details of the pipeline, see [the section corresponding

to workflows in *fMRIPrep*'s documentation](https://fmriprep.readthedocs.io/en/latest/workflows.html "FMRIPrep's documentation").

\### Copyright Waiver

The above boilerplate text was automatically generated by fMRIPrep

with the express intention that users should copy and paste this

text into their manuscripts *unchanged*.

It is released under the [CC0](https://creativecommons.org/publicdomain/zero/1.0/) license.

**Supplemental Tables and Figures**

**Supplementary Table 1. Summary statistics of voxel-wise whole-brain functional connectivity results for the CA1**

| Contrast | Seed | Area | Peak Z | x | y | z | *p*_FDR_  value | voxels | |
| --- | --- | --- | --- | --- | --- | --- | --- | --- | --- |
| CHR-P placebo > HC | | | | | | | | | |
|  | Right CA1 | Right hippocampus | 5.65 | 38 | -20 | -18 | <0.001 | | 6820 |
|  |  | Right inferior temporal gyrus | 4.79 | 48 | -30 | -18 |  |  |  |
|  |  | Right medial temporal gyrus | 4.6 | 50 | -30 | -10 |  |  |  |
|  |  | Right insula | 4.57 | 34 | -14 | -4 |  |  |  |
| CHR-P diazepam > HC | | | | | | | | | |
|  | Right CA1 | Right parahippocampal gyrus | 5.69 | 34 | -28 | -16 | <0.001 | | 5893 |
|  |  | Right hippocampus | 5.67 | 34 | -18 | -20 |  |  |  |
|  |  | Right inferior temporal gyrus | 4.69 | 34 | -8 | -40 |  |  |  |
|  |  | Right fusiform gyrus | 4.68 | 60 | -42 | -14 |  |  |  |
|  |  | Right angular gyrus | 4.86 | 42 | -76 | 30 | <0.001 | | 1545 |
|  |  | Right interparietal sulcus | 3.49 | 16 | -72 | 50 |  |  |  |
| HC > CHR-P placebo | | | | | | | | | |
|  | Right CA1 | Left hippocampus | 5.7 | -32 | -22 | -16 | <0.001 | | 1993 |
|  |  | Left medial temporal gyrus | 4.13 | -58 | -34 | -16 |  |  |  |
|  |  | Right mPFC | 4.37 | 10 | 48 | 16 | 0.040 | | 767 |
|  |  | Left mPFC | 3.73 | -10 | 40 | 30 |  |  |  |
|  |  | Left dorsal ACC | 3.6 | -8 | 30 | 22 |  |  |  |
|  |  | Right dorsal ACC | 3.21 | 14 | 30 | 22 |  |  |  |
|  |  | Left PCC | 3.47 | -4 | -46 | 38 | 0.048 | | 734 |
|  |  | Right PCC | 2.73 | 2 | -22 | 42 |  |  |  |
| HC > CHR-P diazepam | | | | | | | | | |
|  | Right CA1 | Left hippocampus | 4.22 | -32 | -22 | -18 | 0.004 | | 1180 |
|  |  | Left parahippocampal gyrus | 3.78 | -26 | -6 | -34 |  |  |  |

*ACC: anterior cingulate cortex; CHR-P: clinical high-risk for psychosis; FDR: false discovery rate; HC: healthy control; NAc: nucleus accumbens; mPFC: medial prefrontal cortex; PCC: posterior cingulate cortex*
